# Age is Associated with Dampened Circadian Patterns of Rest and Activity: The Study of Muscle, Mobility and Aging (SOMMA)

**DOI:** 10.1101/2023.11.11.23298422

**Authors:** Melissa L. Erickson, Terri L. Blackwell, Theresa Mau, Peggy M. Cawthon, Nancy W. Glynn, Yujia (Susanna) Qiao, Steven R. Cummings, Paul M. Coen, Nancy E. Lane, Stephen B. Kritchevsky, Anne B. Newman, Samaneh Farsijani, Karyn A. Esser

## Abstract

**Background:** Aging is associated with declines in circadian functions. The effects of aging on circadian patterns of behavior are insufficiently described. We characterized age-specific features of rest-activity rhythms (RAR) in community dwelling older adults, both overall, and in relation, to sociodemographic characteristics.

**Methods:** We analyzed baseline assessments of older adults with wrist-worn free-living wrist-worn actigraphy data (N=820, Age=76.4 yrs, 58.2% women) participating in the Study of Muscle, Mobility and Aging (SOMMA). We applied an extension to the traditional cosine curve to map RAR to activity data, calculating the parameters: rhythmic strength (amplitude); robustness (pseudo-F statistic); and timing of peak activity (acrophase). We also used function principal component analysis to determine 4 components describing underlying patterns of activity accounting for RAR variance. Linear models were used to examine associations between RAR and sociodemographic variables.

**Results:** Age was associated with several metrics of dampened RAR; women had stronger and more robust RAR metrics vs. men (all *P* < 0.05). Total activity (56%) and time of activity (20%) accounted for most the RAR variance. Compared to the latest decile of acrophase, those in the earliest decile had higher average amplitude (*P* <0.001). Compared to the latest decile of acroaphase, those is the earliest and midrange categories had more total activity (*P*=0.02). RAR was associated with some sociodemographic variables.

**Conclusions:** Older age was associated with dampened circadian behavior; and behaviors were sexually dimorphic. We identified a behavioral phenotype characterized by early time-of-day of peak activity, high rhythmic amplitude, and more total activity.

## Introduction

Aging is characterized by declines in physical function and mobility. The determinants of these changes are still under investigation. Numerous aging biological processes have been linked to circadian timing, patterns, or rhythms and, thus, the role of circadian biology in age-related changes is now being considered(1). Circadian rhythms are approximate 24hr patterns in behavior and physiology that are regulated by molecular clock mechanisms found in virtually all cells in the body. Endogenous circadian clocks confer benefit to an organism by supporting homeostasis and resilience, and this ultimately promotes longevity and healthy aging(2–4). Mounting evidence suggests that aging itself is characterized by weakened circadian functions(5, 6). In addition, there is a growing interest in linking circadian timing to interventions for healthy aging, including diet(7) and physical activity(8). Nonetheless, there is a need to first establish the fundamental relation between aging and circadian biology.

One observable aspect of circadian biology is the repeated, rhythmic change in rest and activity behaviors. These behavioral circadian patterns are measurable in humans, in free-living settings, with wearable activity monitors(9). Specifically, rest-activity data obtained from such monitors worn for several consecutive days, can be mathematically assessed for a daily circadian rhythm; and the shape of these rhythmic patterns may reveal insight into health and disease status. For example, a remarkably consistent observation across numerous cohort studies is that a dampened rhythmic amplitude is associated with age-related chronic conditions and pathologies, including changes in cognitive functioning, signs of Alzheimer’s disease, fatigue, markers of inflammation, reduced cardiometabolic and bone health, and even mortality (10–20). While these relationships between altered rest-activity rhythms and disease outcomes are striking, what remains unaddressed is the impact of aging itself on rest-activity patterns.

In addition to the features of rest-activity patterns, the time-of-day in which activity occurs is gaining attention as a new parameter of physical activity that is important to health. Studies have reported associations between times of day when activity is performed (e.g. morning, afternoon, or evening) with outcomes that are relevant for age-related chronic diseases, such as obesity, metabolic function, type 2 diabetes, cardiovascular risk, and all-cause mortality(21–25). These findings support an emerging concept of circadian timing of physical activity for health benefit. The circadian patterns of rest and activity in the context of the 24h day-cycle cycle, and whether this relates to healthy aging is unknown. The Study of Muscle, Mobility and Aging (SOMMA) offers opportunity in this regard, enabling large-scale behavior phenotyping of rest-activity rhythms, as well as determination of the temporal distribution of activity in a cohort of older adults (70 to 85+ yrs), free of life-threatening illnesses, did not suffer from mobility disability, and inclusive of men and women(26), which has not been done previously.

The purpose of this study was to determine age-specific features of circadian patterns of rest and activity behavior, assessed with wearable activity trackers, in a cohort of community dwelling older adults in the SOMMA cohort(26). In addition to rhythmic parameters, the temporal distribution of physical activity across the 24h day was also characterized. Finally, associations between parameters of rest-activity-rhythms and demographic variables were examined.

## Methods

### Study Cohort and Design

From April 2019 to December 2021, participants aged 70 and older were recruited from 2 clinical sites—the University of Pittsburgh and Wake Forest University School of Medicine for the Study of Muscle, Mobility and Aging (SOMMA) ((https://sommaonline.ucsf.edu). The unique cohort study design of SOMMA has been previously described elsewhere (26). Briefly, individuals were eligible to participate if they were 70 years old or older, willing and able to complete a skeletal muscle biopsy and undergo magnetic resonance (MR). Individuals were excluded if they reported an inability to walk one-quarter of a mile or climb a flight of stairs; had body mass index (BMI) ≥40 kg/m^2^; had an active malignancy or dementia; or any medical contraindication to biopsy or MR. Finally, participants must have been able to complete the 400-meter walk; those who appeared as they might not be able to complete the 400-meter walk at the in-person screening visit completed a short distance walk (4 meters) to ensure their walking speed as ≥0.6 m/s. SOMMA was approved by the Western IRB-Copernicus Group (WCG) Institutional Review Board (WCGIRB, study number 20180764). All participants provided written informed consent. This current study used baseline SOMMA data for cross-sectional assessments.

### Demographic Variables

Data collected included age based on self-reported date of birth, self-reported gender, and race and self-reported ethnicity based on current census categories. Data on work schedule, education level and finances were gathered. Data on behavior and lifestyle were collected (e.g., smoking status, marital status), self-reported health status and medical history. Multimorbidity was classified using a modification to the Rochester Epidemiology Project multimorbidity scale (0–13) (27). Height was measured by stadiometers and weight by balance beam or digital scales. Body mass index (BMI) was then calculated as weight (kg)/height (m^2^).

### Actigraphy

Actigraphy data was collected using the ActiGraph GT9X (ActiGraph, Pensacola, FL), which has a 3-axis accelerometer with a sampling rate of 80 Hertz. ActiGraph GT9X is a watch-like device placed on a participant’s nondominant wrist in person at a clinic visit. Participants were asked to wear the Actigraph continuously for 7 days(28). Data were processed in one-minute epochs (activity counts/minute) and scored using ActiGraph Corp’s ActiLife Software. The first day of wear was excluded from these analyses, as participants were required to do a number of physical performance tests during their clinic visit and the activity level may not be representative of their usual activity patterns. Sleep diaries were used to aid in setting intervals for when the participants were in bed trying to sleep. Nonwear time was determined by a combination of an off-wrist detector in the device, a nonwear algorithm, and review by an actigraphy data scorer(28, 29). Nonwear times were set to missing. The Cole-Kripke sleep scoring algorithm was used to determine sleep from wake(30). Total sleep time during the in-bed interval was averaged over all nights of wear, to obtain a more representative characterization of usual sleep patterns. Total activity count per 24-hour day was also averaged over all days to get an estimate of overall activity level.

### Rest-Activity Rhythm Parameters

The activity data gathered was used to calculate both parametric and non-parametric RAR variables. The parametric approach assumes the activity data has an underlying distribution similar to the cosine curve. The nonparametric approach does not assume RAR fit to a cosine wave *a priori* but rather fits to regular pattern of activity.

#### Parametric Approach

A 5-parameter extension to the traditional 24-hr cosine curve was used to map the RAR to activity data. This extension allows for a more squared-shape wave than a cosine curve, as often observed with activity data (31). The RAR parameters include the following: amplitude, which is an indicator of the strength of the rhythm, calculated as the peak to nadir difference in activity (units of activity [counts/min]); midline (midpoint between the rhythmic maximum and minimum), estimating statistic of rhythm (mesor), which is the mean level of activity (units of activity [counts/min]); robustness of the RAR, or pseudo-F statistic for goodness of extended cosine fit, with higher values indicate stronger rhythms; and acrophase, which is the timing of peak activity of the fitted curve, measured as time of day (portions of hours).

#### Non-Parametric Approach

Inter-day stability (IS), which describes day-to-day stability of RAR (range 0 to 1); and intra-daily variability (IV) which describes fragmentation across 24h ranges (range 0 to 2); the average activity level of the most active consecutive 10-hour period (M10); the average activity of the least active consecutive 5-hour period (L5); relative amplitude (RA), the difference in activity between M10 and L5 in the average 24-hour pattern, normalized by their sum, with higher RA reflecting relatively lower activity during the night and greater activity when awake (32, 33).

#### Functional Principal Component Analysis (fPCA)

We also used fPCA to describe underlying pattens of activity, as this analytical approach does not rely on *a priori* assumptions about the activity shape. Participant data was fit with a nine-Fourier-based function. fPCA was then used to derive the top four components determined as these typically explain the majority of the variance, and an eigenvalue was assigned for each of the four components and each participant(34, 35).

#### Temporal Distribution of Physical Activity

The average of activity level across all participants by clock time were plotted, stratified by acrophase category. Participants were categorized as having early timing if they fell within the lowest decile of acrophase, midrange for those 10% of participants around the median value, and late timing as those in the highest decile of acrophase.

## Statistical Analysis

Cohort characteristics were categorized and described using proportions (N% of). RAR parameters were described using means and standard deviations. Associations of each characteristic with the RAR parameters was examined using linear regression models, with results presented as adjusted means and their 95% confidence intervals. For characteristics with more than 2 categories, tests for a linear trend across categories were performed by including each characteristic (ordinal variable) as an independent variable in models. Tests were also performed comparing categories to the reference. Minimally adjusted models included the characteristic and an adjustment for clinic site. Multivariable adjusted models included clinic site and all characteristics examined in the same model, to determine if adjustment for other characteristics attenuated any associations observed.

We explored differences in associations by sex by performing formal tests for interaction with sex and each characteristic with linear regression models that included clinic site, the characteristic, sex, and a term for sex*characteristic.

Total activity level across categories of acrophase used to describe the temporal distribution of activity were compared using t-tests, comparing the participants in the midrange group to those in the lowest and highest decile of acrophase. In addition, area-under-the-curve (AUC) for the graphical representation of average activity stratified by category of acrophase was calculated using the trapezoidal rule.

All significance levels reported were two-sided and all analyses were conducted using SAS version 9.4 (SAS Institute Inc, Cary, NC).

## Results

### Participants

Of the 879 participants enrolled in SOMMA, our analytic subset consists of 820 participants with actigraphy data. Some participants (n=59) missing or excluded were due to several reasons; either the participant wore the device but there was a malfunction with the datafile (n=33), no device was available (n=12), the participant refused (n=1), the participant was unable (n=1), the actigraphy file did not have activity data in the correct format (n=9) or had too little data collected (n=3). The 820 men (41.8%) and women (58.2%) were on average 76.4 years old, had a BMI of 27.6 kg/m^2^, and mostly identified as White (85.0%). Most (62.0%) graduated from college and about half were in a married-like relationship. Most (61.6%) reported very good or excellent health compared to others their age and 83.3% reported a history of one or more of the 13 medical conditions in the multimorbidity index. Most said their finances met their needs very well (64.1%) and some (39.4%) reported having a regular work or volunteer schedule (Table 1). Only 20% of participants reported regularly waking with an alarm, and remaining 80% had different self-wake behaviors, potentially indicating that they were not constrained by scheduled requirements. The participants on average slept 6 hours, 51 minutes ± 61 minutes.

**Table 1:** Associations of descriptive variables with rest-activity rhythm parameters. Site adjusted means (95% CI).

| Descriptive | N (%) In Category | Amplitude (counts/min) | Parametric |  |  | Nonparametric |  |
| --- | --- | --- | --- | --- | --- | --- | --- |
|  |  |  | Mesor (counts/min) | Acrophase (portions of hours) | Pseudo-F value | Interdaily Stability (range 0-1) | Intradaily Variability (range 0-2) |
| **Unadjusted mean ± SD** | | 2183.5 ± 1143.4 | 1307 ± 618.0 | 14.31 ± 1.5 | 670.1 ± 302.4 | 0.58 ± 0.12 | 0.89 ± 0.22 |
| **Age, years** |  |  |  |  |  |  |  |
| 70-74 (reference) | 377 (46.0) | 2328.7 (2214.3, 2443.2) | 1377.6 (1315.5, 1439.7) | 14.3 (14.1, 14.4) | 674.1 (643.6, 704.6) | 0.58 (0.56, 0.59) | 0.88 (0.86, 0.90) |
| 75-79 | 252 (31.7) | 2170.8 (2030.9, 2310.8) | 1289.7 (1213.8, 1365.6) | 14.4 (14.2, 14.6) | 692.3 (655.0, 729.6) | 0.59 (0.57, 0.60) | 0.87 (0.84, 0.89) |
| 80-84 | 125 (15.2) | 1993.4 (1794.5, 2192.2)** | 1218.7 (1110.8, 1326.5)* | 14.2 (13.9, 14.5) | 624.4 (571.3, 677.4) | 0.57 (0.55, 0.59) | 0.96 (0.92, 1.00)*** |
| 85+ | 66 (8.1) | 1762.7 (1489.2, 2036.2)** | 1137.2 (988.8, 1285.5)** | 14.3 (14.0, 14.7) | 649.5 (576.6, 722.5) | 0.58 (0.55, .061) | 0.94 (0.89, 1.00)* |
| *P*-trend |  | <0.001 | <0.001 | 0.96 | 0.23 | 0.83 | 0.001 |
| **Sex** |  |  |  |  |  |  |  |
| Men | 343 (41.8) | 2119.6 (1998.5, 2240.6) | 1286.6 (1221.2, 1352.1) | 14.2 (14.0, 14.4) | 605.0 (573.5, 636.5) | 0.56 (0.55, 0.57) | 0.92 (0.90, 0.95) |
| Women | 477 (58.2) | 2229.5 (2126.9, 2332.1) | 1321.7 (1266.1, 1377.2) | 14.4 (14.3, 14.5) | 717.0 (690.3, 743.7) | 0.59 (0.58, 0.60) | 0.87 (0.85, 0.89) |
| *P*-value |  | 0.17 | 0.42 | 0.07 | <0.001 | <0.001 | <0.001 |
| **Race** |  |  |  |  |  |  |  |
| White | 697 (85.0), | 2213.0 (2128.2, 2297.9) | 1313.6 (1267.7, 1359.5) | 14.3 (14.2, 14.4) | 678.4 (656.0, 700.9) | 0.59 (0.58, 0.60) | 0.90 (0.88, 0.92) |
| Non-White | 123 (15.0) | 2016.3 (1814.3, 2218.2) | 1269.6 (1160.3, 1379.0) | 14.5 (14.2, 14.7) | 623.3 (569.9, 676.6) | 0.54 (0.52, 0.56) | 0.85 (0.81, 0.89) |
| *P*-trend |  | 0.08 | 0.47 | 0.22 | 0.06 | <0.001 | 0.03 |
| **Education Level** |  |  |  |  |  |  |  |
| High school or less or other | 121 (14.9) | 2235.1 (2030.2, 2440.1) | 1366.6 (1255.9, 1477.3) | 14.3 (14.0, 14.5) | 700.6 (646.8, 754.5) | 0.57 (0.55, 0.60) | 0.86 (0.83, 0.90)* |
| Some college | 188 (23.2) | 2146.7 (1982.0, 2311.4) | 1265.1 (1176.2, 1354.1) | 14.5 (14.3, 14.7) | 709.4 (666.1, 752.7)* | 0.59 (0.57, 0.60) | 0.85 (0.81, 0.88)*** |
| College Graduate | 209 (25.7) | 2164.0 (2007.9, 2320.2) | 1320.6 (1236.3, 1405.0) | 14.1 (13.9, 14.3) | 645.0 (604.0, 686.1) | 0.57 (0.55, 0.59) | 0.92 (0.89, 0.95) |
| Post College Graduate (reference) | 294 (36.2) | 2206.8 (2074.9, 2338.8) | 1302.8 (1231.6, 1374.1) | 14.3 (14.1, 14.5) | 650.1 (615.4, 684.8) | 0.58 (0.57, 0.60) | 0.92 (0.89, 0.94) |
| *P*-trend |  | 0.97 | 0.69 | 0.54 | 0.03 | 0.77 | <0.001 |
| **How well money takes care of needs at end of month** |  |  |  |  |  |  |  |
| Refused/Poorly | 41 (5.0) | 1891.4 (1540.8, 2241.9) | 1165.1 (975.5, 1354.7) | 14.8 (14.4, 15.3)* | 590.8 (498.6, 683.1)* | 0.54 (0.50, 0.58)** | 0.93 (0.86, 1.00) |
| Fairly well | 252 (30.9) | 2138.1 (1996.4, 2279.8) | 1307.4 (1230.7, 1384.0) | 14.5 (14.3, 14.7)* | 630.0 (592.7, 667.2)** | 0.55 (0.54, 0.57)*** | 0.88 (0.86, 0.91) |
| Very well (reference) | 522 (64.1) | 2231.6 (2133.3, 2329.9) | 1320.0 (1266.9, 1373.2) | 14.2 (14.1, 14.3) | 696.1 (670.2, 721.9) | 0.60 (0.58, 0.61) | 0.90 (0.88, 0.91) |
| *P*-trend |  | 0.06 | 0.25 | 0.001 | 0.001 | <0.001 | 0.97 |
| **Work or volunteer schedule** |  |  |  |  |  |  |  |
| No regular schedule | 492 (60.6) | 2101.0 (2008.8, 2211.1) | 1279.0 (1224.2, 1333.8) | 14.3 (14.2, 14.5) | 665.5 (638.7, 692.3) | 0.58 (0.57, 0.59) | 0.90 (0.88, 0.92) |
| Regular schedule | 320 (39.4) | 2304.1 (2178.7, 2429.5) | 1352.0 (1284.1, 1420.0) | 14.3 (14.1, 14.5) | 678.8 (645.6, 712.1) | 0.58 (0.56, 0.59) | 0.88 (0.86, 0.90) |
| *P*-trend |  | 0.02 | 0.10 | 0.69 | 0.54 | 0.47 | 0.14 |
| **Marital status** |  |  |  |  |  |  |  |
| Married/in married-like relationship | 418 (51.2) | 2306.0 (2196.6, 2415.4) | 1367.6 (1308.4, 1426.8) | 14.2 (14.0, 14.3) | 694.9 (665.9, 723.9) | 0.60 (0.58, 0.61) | 0.90 (0.88, 0.93) |
| Unmarried | 398 (48.8) | 2052.8 (1940.7, 2164.9) | 1242.5 (1181.9, 1303.2) | 14.4 (14.3, 14.6) | 642.3 (612.7, 672.0) | 0.56 (0.55, 0.57) | 0.88 (0.86, 0.90) |
| *P*-trend |  | 0.002 | 0.004 | 0.01 | 0.01 | <0.001 | 0.17 |
| **Self-reported health status** |  |  |  |  |  |  |  |
| Good or fair | 313 (38.5) | 2054.8 (1928.0, 2181.6) | 1245.3 (1176.7, 1313.9) | 14.5 (14.3, 14.7) | 650.0 (616.5, 683.6) | 0.57 (0.56, 0.58) | 0.89 (0.87, 0.92) |
| Excellent or very good | 501 (61.6) | 2264.1 (2163.9, 2364.3) | 1345.5 (1291.3, 1399.7) | 14.19 (14.1, 14.3) | 682.0 (655.5, 708.5) | 0.59 (0.57, 0.60) | 0.89 (0.88, 0.91) |
| *P*-trend |  | 0.01 | 0.03 | 0.003 | 0.14 | 0.07 | 0.82 |
| **Number of multimorbidities (0-13)\*\*\*** |  |  |  |  |  |  |  |
| None (reference) | 134 (16.7) | 2242.9 (2048.4, 2437.3) | 1343.2 (1238.4, 1448.1) | 14.2 (13.9, 14.4) | 701.80 (650.63, 752.97) | 0.59 (0.57, 0.61) | 0.88 (0.84, 0.91) |
| 1 | 284 (35.3) | 2252.1 (2118.5, 2385.7) | 1336.7 (1264.7, 1408.8) | 14.2 (14.0, 14.4) | 676.62 (641.46, 711.77) | 0.58 (0.56, 0.59) | 0.91 (0.88, 0.94) |
| 2 | 246 (30.6) | 2141.2 (1997.7, 2284.8) | 1283.9 (1206.6, 1361.3) | 14.5 (14.3, 14.7)* | 645.28 (607.51, 683.04) | 0.57 (0.56, 0.59) | 0.88 (0.86, 0.91) |
| 3+ | 140 (17.4) | 2083.9 (1893.5, 2274.4) | 1259.0 (1156.4, 1361.7) | 14.4 (14.1, 14.6) | 675.86 (625.76, 725.97) | 0.59 (0.57, 0.61) | 0.89 (0.86, 0.93) |
| *P*-trend |  | 0.13 | 0.15 | 0.07 | 0.27 | 0.81 | 0.98 |
| **Smoking status** |  |  |  |  |  |  |  |
| Never smoked | 457 (56.1) | 2184.8 (2079.5, 2290.2) | 1303.0 (1246.1, 1359.9) | 14.4 (14.2, 14.5) | 663.65 (635.84, 691.45) | 0.57 (0.56, 0.58) | 0.90 (0.88, 0.92) |
| Current or past smoker | 358 (43.9) | 2184.6 (2065.5, 2303.6) | 1314.1 (1249.8, 1378.4) | 14.3 (14.1, 14.4) | 677.54 (646.11, 708.97) | 0.59 (0.57, 0.60) | 0.89 (0.87, 0.91) |
| *P*-trend |  | 1.0 | 0.80 | 0.39 | 0.52 | 0.15 | 0.64 |

All models are adjusted by clinic site (RAR parameter~clinic site + one descriptive characteristic in separate models).
For predictors with >2 categories, a *P*-trend was calculated, looking for a linear trend across the categories. Categories were also compared to the reference category.
The symbols represent the *P* -value for the comparison of the category to the reference category.
Symbols: \*= *P*-value<0.05; \*\*= *P*-value<0.01; \*\*\* *P*-value<0.001

### Parametric and Non-Parametric Rest-Activity Rhythmic Parameters

Representative examples of rest-activity rhythms are shown in Figure 1. On average, participants wore the ActiGraph for 8 ± 0.8, 24-hr periods. The average acrophase was at 2:19 PM. The average IS and IV were 0.58 and 0.59, respectively (Table 1, Supplemental Figure S1).

**Figure 1:**
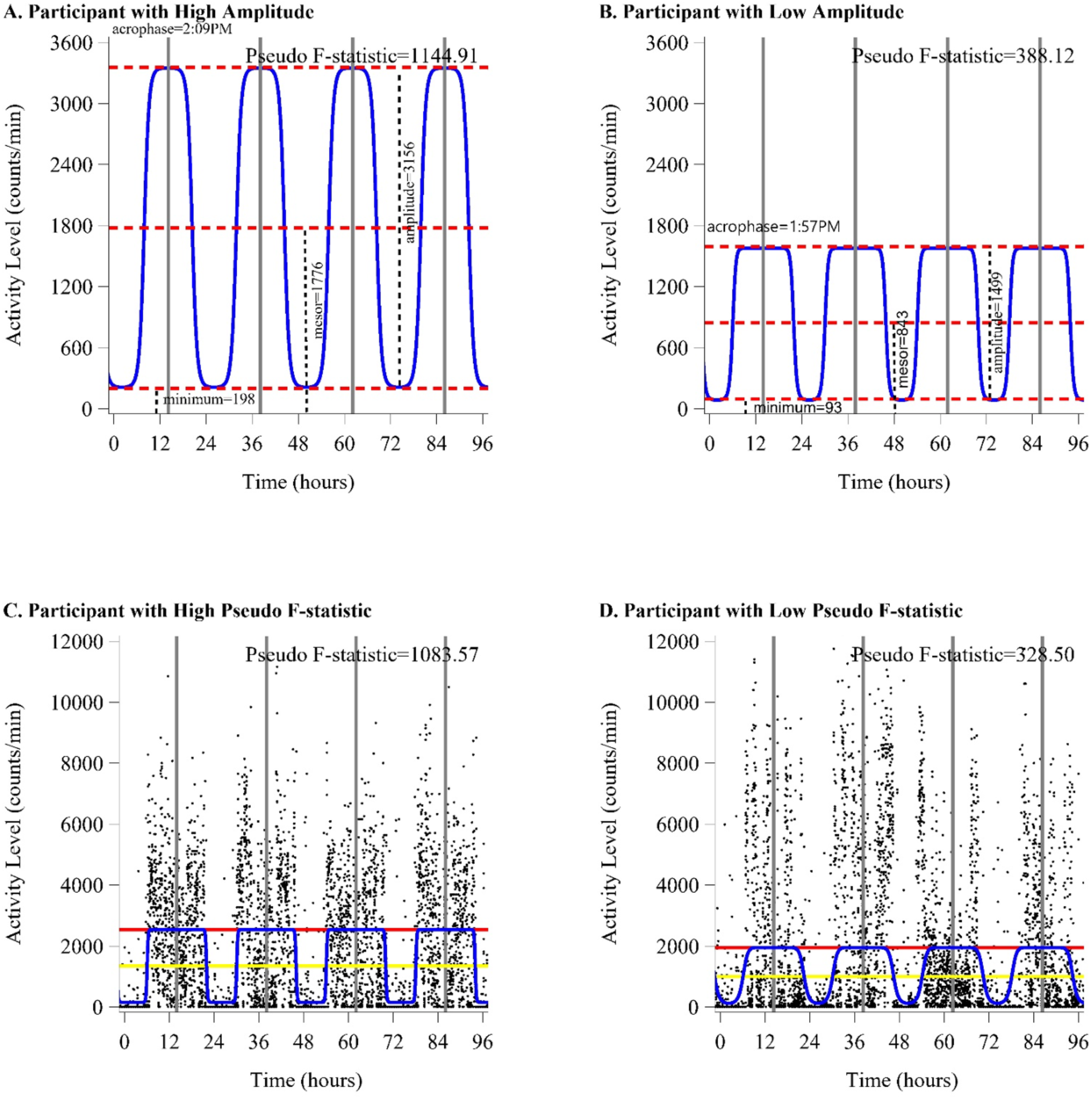
Representative examples of rest-activity rhythm profiles demonstrating differences in rhythmic amplitude and rhythmic strength in community-dwelling men and women 70 and older: the SOMMA Cohort. Comparison of representative rest-activity rhythm plots of individual participants from the highest 10^th^ percentile of amplitude (Panel A) versus lowest 10^th^ percentile of amplitude (Panel B). Amplitude, minimum, and mesor are labeled with red dashed line. Acrophase (time of peak activity) is shown with a gray bar. Comparison of representative rest-activity rhythms of individual participants from the lowest decile values for pseudo F-statistic (Panel C) versus the highest decile values for pseudo F-statistic (Panel D) to graphically illustrate stronger rhythmic strength with clear sleep-wake patterns versus weaker rhythmic strength with less distinct sleep-wake patterns. Mesor (yellow line), amplitude (red line), fitted curve (blue line) and acrophase (gray bar) are labeled.

### Function Principal Component Analysis

The four components of the fPCA analysis explained 91% of the variance in the activity data. The first component primarily described overall activity level (fPCA1: 56% of the variance), the second component primarily described timing of activity (fPCA2: 20% of the variance), the third component primarily described a lower level of midday activity (fPCA3: 9% of the variance), and the fourth component primarily differentiated between a morning activity peak and an afternoon peak (fPCA4: 6% of the variance). Figure 2 shows the plots of activity level for the average of the cohort, those with positive eigenvalues and those with negative eigenvalues for each of the 4 fPCA.

**Figure 2:**
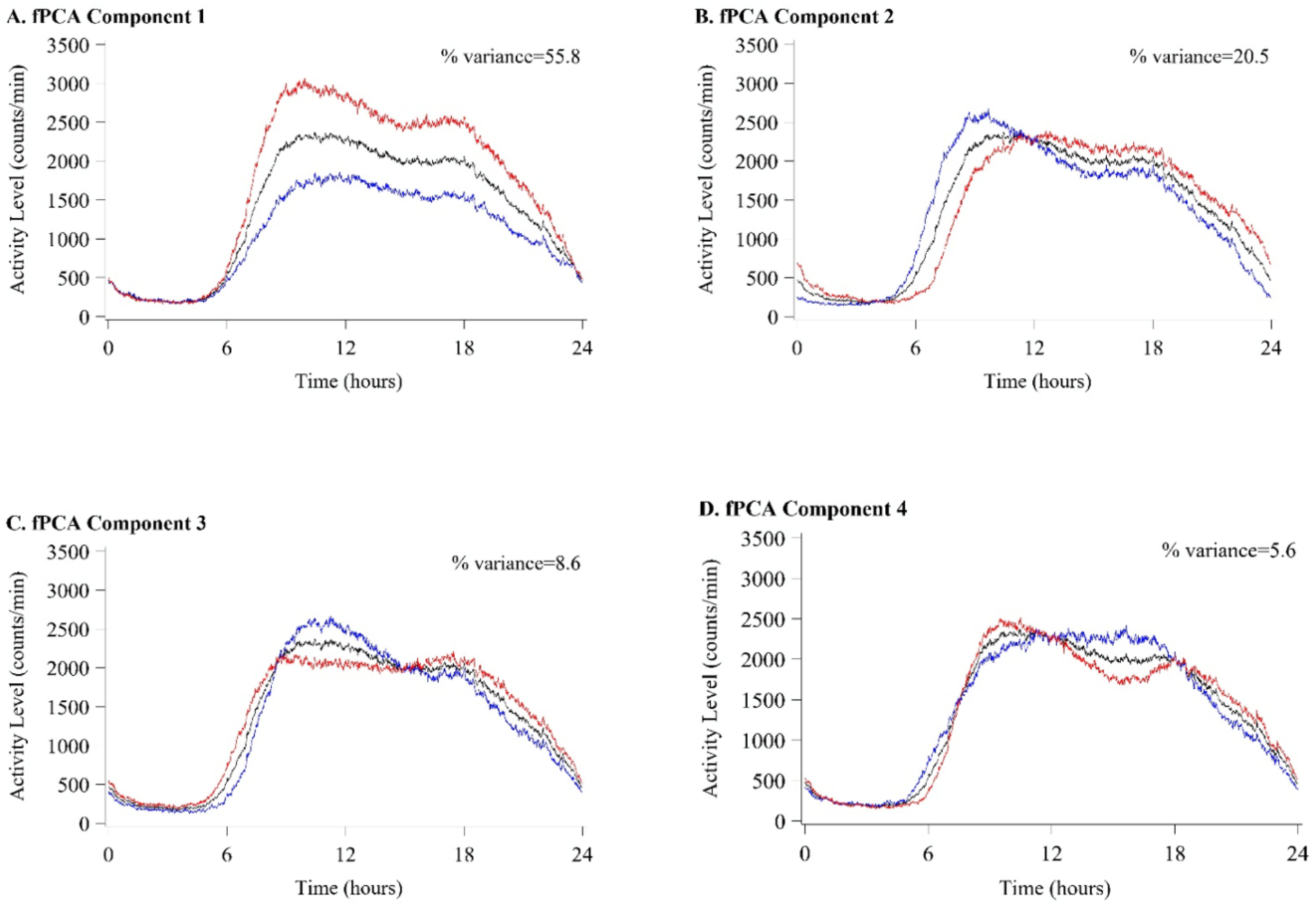
Four components of functional principal component analysis (fPCA). The average pattern of activity for all participants (black line); average pattern of activity in participants with the eigenvalue of positive fPCA scores (red line); average pattern of activity in participants with the eigenvalue of negative fPCA scores (blue line). fPCA1 represents high and low overall activity explaining 55.8% of variance (Panel A). fPCA2 represents later activity timing (positive eigenvalues) and earlier (negative eigenvalues) activity timing (Panel B) explaining 20.5% of variance. fPCA3 represents longer, biphasic (low eigenvalues) and shorter, more monophasic (high eigenvalues), activity patterns explaining 8.6% of variance (Panel C). fPCA4 represents morning (high eigenvalues) and evening (low eigenvalues) peaks in activity explaining 5.6% of variance (Panel D).

### Associations Between RAR Parameters

Measures that are primarily related to activity level from the 3 approaches of defining RAR were highly correlated to each other (r>0.50 for amplitude, mesor, M10, fPCA component 1; Supplemental Figure 1). Acrophase and fPCA component 2, both measures of timing were correlated at r=0.75. Measures of rhythm robustness or fragmentation were also highly correlated (abs(r)>0.64 for pseudo F-statistic, IS, IV; Supplemental Figure S2).

### Associations of RAR Parameters with Demographic Variables

In models adjusted for clinic site alone, age was primarily related to parameters that are driven by activity level and strength of rhythm, in which younger participants had higher average values of amplitude, mesor, M10, and fPCA1; lower values of IV. Sex was primarily related to the strength of patterns of activity (pseudo f-statistic, IS, IV, M10, RA, fPCA1, fPCA2). Figure 3 shows sex differences in parametric and non-parametric parameters. Race was not related to any shape-based parameters, but was related to nonparametric measures, with those identifying as White having higher stability (IS), lower variability (IV) and lower L5 (Table 1, Supplemental Table 1).

**Figure 3:**
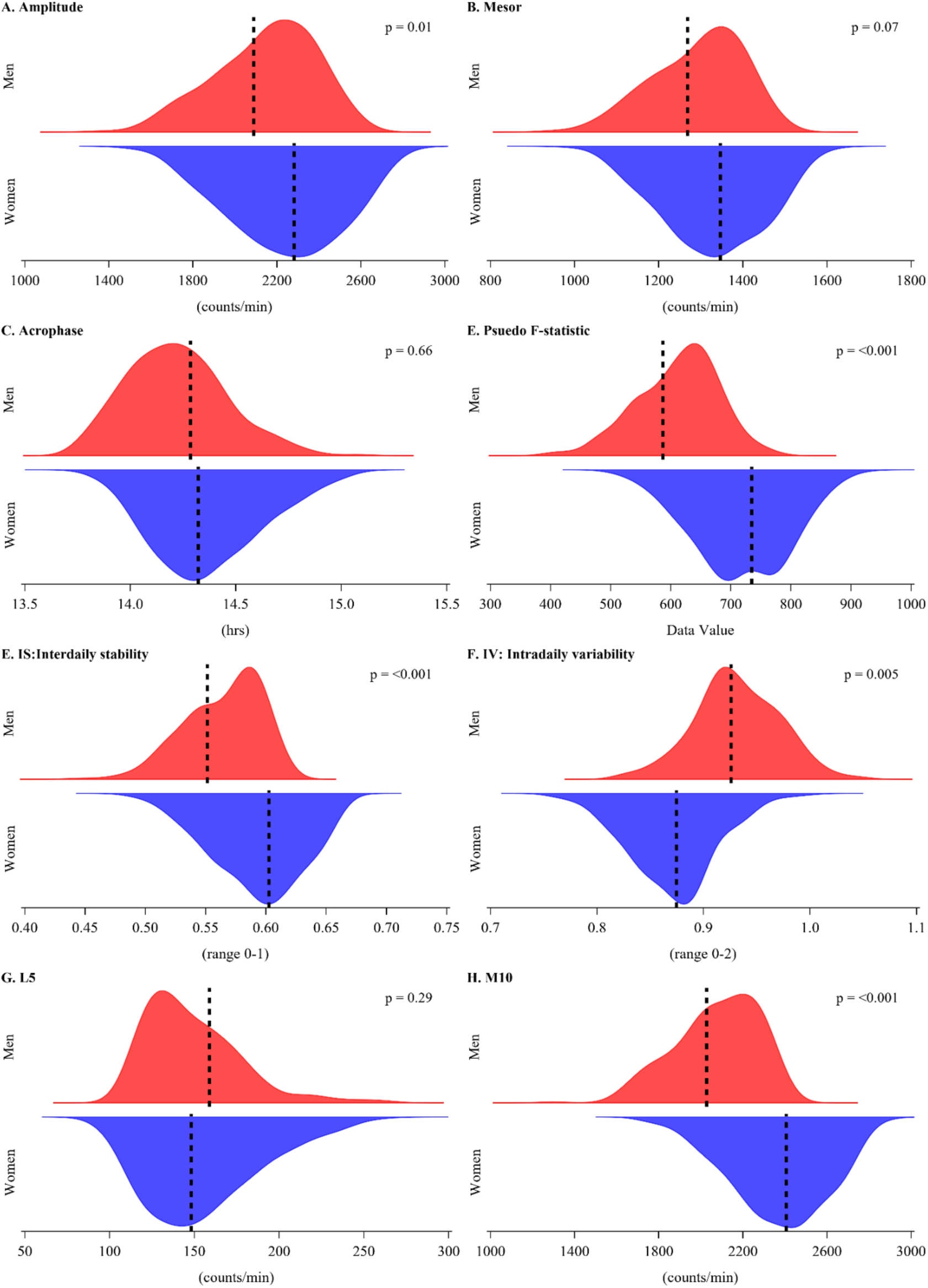
Older community-dwelling. women have higher rhythmic amplitude and rhythmic strength compared to male counterparts. Kernel density plots of multiple adjusted predicted values shown separately by men (red) and women (blue) for parametric and non-parametric parameters, including rhythmic amplitude (Panel A), mesor (Panel B), acrophase (Panel C), Psuedo F-statistic (Panel D), interdaily stability (Panel E), intradaily variability (Panel F), L5 (Panel G) and M10 (Panel H). Dashed lines represent adjusted means. Model adjusted for clinic site plus all characteristics examined. *P*-values represents comparison between sexes.

The most consistent association seen was that of marital status and RAR (Table 1, Supplemental Table 1). Being in a married-like relationship was associated with more robust rhythms as seen by the parametric parameters, more stability of activity (IS), lower levels of L5, implying more consolidated sleep, and higher M10 (more active while out of bed). Those with higher education level had less strength of rhythm (pseudo f-statistic, IV, L5, RA). Financial situation was related to timing of activity and strength of rhythm (acrophase and pseudo f-statistic), and most nonparametric measures (IS, L5, M10, RA, fPCA1, fPCA2). The associations of work were primarily activity level based (amplitude, M10, fPCA1). Reporting poor/good health status was primarily related to lower average activity levels. There were no associations observed between smoking or the multimorbidity index and RAR parameters.

Associations seen in the site-adjusted models remained statistically significant for most demographic variables after combining all demographic variables in one model, with some attenuation of effect size (Supplement Tables 2A, B, C). The demographic variables most affected by adjustment for other variables examined were work schedule and self-reported health status.

There were very few significant interactions between sex and other demographic variables. There were no significant interactions of sex seen with age, race, education, financial security, self-reported health status or smoking (*P* > 0.05). The interaction of sex with the multimorbidity index was significant for amplitude and fPCA1 (*P* < 0.05), but associations were not statistically significant after stratification by sex.

### Temporal Distribution of Activity

As described above, acrophase is the time of day of peak activity. Figure 4 shows plots of the average of activity across the day for all participants by category of acrophase. Those with the earlier acrophase (<12:43 PM) had the highest peak activity and a sharp decline later in the evening. Those with the latest acrophase (>3:55 PM) had more activity in the evening (11PM to 2 AM). Compared to the latest decile of acrophase, those in the earliest decile of acrophase had a 70% higher average amplitude (*P* <0.001).

**Figure 4:**
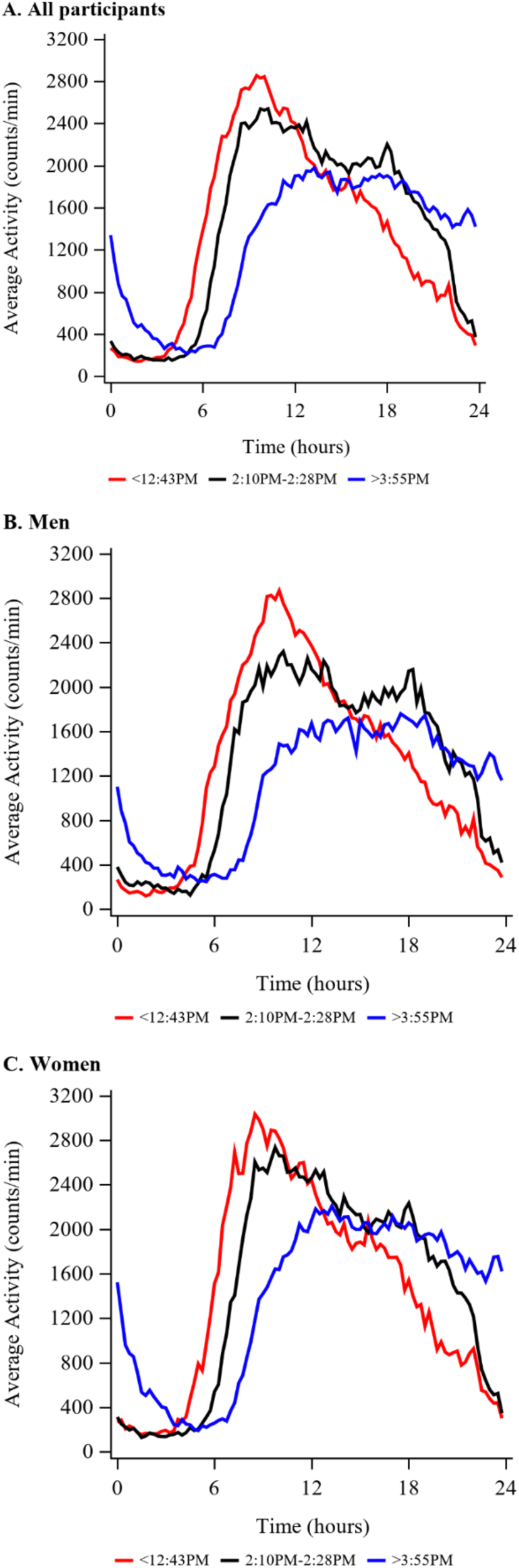
Temporal distribution of average activity across 24h by category of acrophase in community-dwelling older adults. Graphical representation of average activity stratified by category of acrophase (lowest decile: <12:43 PM, red line, middle decile (45-55 percentile): 2:10-2:28 PM, black line; upper decile: >3:55 PM, blue line) over all participants (Panel A), and also separated by men (Panel B) and women (Panel C).

The AUC of the plots show that average activity is similar for those in the earliest and midrange acrophase categories (Figure 4, Panel A: 32648.15 vs. 33752.31); whereas those in the latest category of activity timing had a lower AUC (30117.93). Women had a higher AUC than men (midrange timing category: 34891.61 vs. 31634.91).

Total activity was compared among the three acrophases. The average activity level of those in the midrange category of acrophase was 203.75 ± 46.67 counts*10,000. Compared to those in the midrange groups of acrophase, on average, those in the earliest acrophase category had a similar 24-hr activity level (197.27 ± 53.78 counts*10,000, *P* =0.41), while those in the latest acrophase category had lower 24-hr activity level compared to those in the midrange group (183.68 ± 62.91 counts*10,000; *P*=0.02).

## Discussion

The primary finding of this study was that older age was associated with several metrics of dampened rest-activity rhythms. This is in agreement with findings from a large cohort study representative of the general population, from the National Health and Nutritional Examination Survey (NHANES), which primarily focused on younger age categories (20-39, 40-59, ≥ 60 yrs) (36). We also observed sexual dimorphism in circadian behavior, in that women had stronger and more robust rest-activity rhythms compared to men. This finding is also consistent with two previous large cohort studies, representative of the general population (NHANES and United Kingdom Biobank) (36, 37). As SOMMA focused on older adults, our observations herein indicate that sex-specific differences in circadian behavior may persist beyond reproductive potential, which has not been previously demonstrated. Despite this sexual dimorphism, there was lower rhythmic strength at higher age in both men and women, perhaps supporting the notion that age is a central determinant of circadian patterns of behavior. Although this was a cross-sectional analysis, our findings of dampened circadian rest-activity rhythms in older adults, suggests that these changes are likely paralleled by age-related declines in function, mobility, and energy. Future studies to disentangle cause and effect are warranted.

Function principal component analysis revealed that the two primary components explained a majority of the variance in activity profiles. The first component was overall activity (56%), in which a higher value corresponds with higher activity throughout the day. The second component was time of activity (20%), which corresponds with activity timing (e.g early vs. later “rises”. These findings are very similar to that observed in NHANES, which reported that variance in activity profiles was also primarily explained by overall activity (50%) and timing of activity (21%)(38). The consistency between SOMMA and NHANES cohorts, which, as noted above focused on different age ranges, suggests that patterns of activity profiles are generally preserved from middle to older age.

To better understand activity patterns within the context of the 24h day-night timescale, we investigated the temporal distribution of physical activity. This analysis yielded new insight, in that those with the earliest time-of-day of peak activity (<12:43 PM) had a higher rhythmic peak; whereas, those with the latest time-of-day of peak activity (>3:55 PM) had a lower rhythmic peak. This is the first time, of which we are aware, to describe this behavior phenotype. There appears to be a relation between time-of-day of activity and total daily activity, as those in the earliest and midrange categories performed more total activity compared to those in the latest category of activity timing. Based on these observations, one might suspect that a strong and robust circadian pattern of activity facilitates the accumulation of more total daily activity. Although speculative, perhaps this is one way in which circadian rhythms enable higher levels of physical activity, which in turn promotes healthy aging.

In addition to age and sex, there were some significant associations with rhythmic parameters and sociodemographic variables. Being in a married-like relationship was associated with stronger and more robust rhythms, higher education was associated with less rhythm strength, and financial situation was associated with timing of activity and rhythm strength.

Previous analyses from NHANES have reported associations between race/ethnicity and rhythmic parameters, which were not replicated herein, and this is mostly likely due to differences in samples sizes of diverse races/ethnicities between study cohorts. However, our current observations provide additional context, in which some sociodemographic variables, in addition to age and sex, are associated with rest-activity patterns in community dwelling older adults.

## Conclusion

We found that age was associated with dampened circadian patterns of rest and activity, and this sheds light on a new temporal dimension by which aging impacts physical activity. In addition, women had stronger and more robust rhythms relative to men counterparts. Given the sex gap in longevity and lifespan(39), it is tempting the speculate that strong and robust rhythms in women confers some type of benefit that promotes resiliency or delays aging. We also observed that those active at earlier times in the 24 hour/day had a higher rhythmic peak and more total activity. This may suggest that a strong and robust circadian rhythm facilitates higher levels of, or greater engagement with, physical activity. This novel and comprehensive characterization of rest-activity rhythms in older, community dwelling adults, free of life-threatening disease, lays new groundwork for future hypothesis testing; indeed, future studies that determine how these rest-activity patterns intertwine with function and mobility are warranted.

## Data Availability

This dataset is available upon formally requesting, accepting clinical data use agreements, and receiving approval at https://sommaonline.ucsf.edu/.

## Supplemental Materials

**Supplemental Table 1A:** Associations of descriptive variables with rest-activity rhythm parameters. Site adjusted means (95% CI).

| Descriptive | N (%) In Category | L5:<br>counts/min | M10:<br>counts/min | Relative<br>amplitude<br>((M10-L5)/(M10+L5)) |
| --- | --- | --- | --- | --- |
| **Unadjusted mean**<br>**± SD** |  | 152.94 ± 114.64 | 2236.69 ± 664.46 | 0.87 ± 0.09 |
| **Age (yrs)** |  |  |  |  |
| 70-74 (reference) | 377 (46) | 156.57 (145.00,<br>168.14) | 2355.46 (2289.88,<br>2421.04) | 0.87 (0.86,<br>0.88) |
| 75-79 | 252 (31) | 147.60 (133.45,<br>161.75) | 2253.12 (2172.93,<br>2333.31) | 0.87 (0.86,<br>0.88) |
| 80-84 | 125 (15) | 142.07 (121.96,<br>162.17) | 1979.76 (1865.82,<br>2093.71)*** | 0.86 (0.85,<br>0.88) |
| 85+ | 66 (8) | 173.13 (145.47,<br>200.78) | 1982.09 (1825.37,<br>2138.80)*** | 0.85 (0.82,<br>0.87) |
| *P*-trend |  | 0.99 | <0.001 | 0.11 |
| **Sex** |  |  |  |  |
| Males | 343 (42) | 149.87 (137.73,<br>162.01) | 2061.65 (1993.03,<br>2130.28) | 0.86 (0.85,<br>0.87) |
| Females | 477 (58) | 155.14 (144.85,<br>165.44) | 2362.55 (2304.36,<br>2420.74) | 0.87 (0.86,<br>0.88) |
| *P*-value |  | 0.52 | <0.001 | 0.03 |
| **Race** |  |  |  |  |
| White | 697 (85) | 143.84 (135.48,<br>152.21) | 2251.35 (2202.04,<br>2300.67) | 0.87 (0.87,<br>0.88) |
| Non-White | 123 (15) | 204.47 (184.56,<br>224.38) | 2153.59 (2036.19,<br>2270.99) | 0.82 (0.80,<br>0.84) |
| *P*-trend |  | <0.001 | 0.13 | <0.001 |
| **Education Level** |  |  |  |  |
| High school or<br>less or other | 121 (14.90) | 178.12 (157.76,<br>198.48)** | 2257.91 (2139.17,<br>2376.65) | 0.85 (0.83,<br>0.86)** |
| Some college | 188 (23.15) | 158.58 (142.22,<br>174.94) | 2292.44 (2197.03,<br>2387.85) | 0.87 (0.85,<br>0.88) |
| College Graduate | 209 (25.74) | 149.98 (134.47,<br>165.49) | 2186.80 (2096.32,<br>2277.28) | 0.86 (0.85,<br>0.87) |
| Post College<br>Graduate<br>(reference) | 294 (36.21) | 140.24 (127.13,<br>153.34) | 2230.53 (2154.08,<br>2306.99) | 0.88 (0.87,<br>0.89) |
| *P*-trend |  | 0.002 | 0.40 | 0.01 |
| **How well money takes care of needs at end of month** |  |  |  |  |
| Refused/Poorly | 41 (5.03) | 184.16 (149.50,<br>218.82)* | 1932.95 (1731.53,<br>2134.37)*** | 0.83 (0.80,<br>0.85)*** |
| Fairly well | 252 (30.92) | 179.31 (165.30,<br>193.32)*** | 2150.34 (2068.94,<br>2231.73)** | 0.84 (0.83,<br>0.85)*** |
| Very well (reference) | 522 (64.05) | 137.63 (127.91, 147.35) | 2303.67 (2247.19, 2360.14) | 0.88 (0.87, 0.89) |
| *P*-trend |  | <0.001 | <0.001 | <0.001 |
| **Work or Volunteer Schedule** |  |  |  |  |
| No regular schedule | 492 (60.59) | 150.51 (140.39, 160.63) | 2193.86 (2135.29, 2252.42) | 0.87 (0.86, 0.87) |
| Regular schedule | 320 (39.41) | 155.88 (143.32, 168.43) | 2303.04 (2230.40, 2375.68) | 0.87 (0.86, 0.88) |
| *P*-trend |  | 0.51 | 0.02 | 0.57 |
| **Marital Status** |  |  |  |  |
| Married/in married-like relationship | 418 (51.23) | 135.66 (124.76, 146.57) | 2298.24 (2234.71, 2361.77) | 0.88 (0.87, 0.89) |
| Unmarried | 398 (48.77) | 171.05 (159.87, 182.22) | 2167.47 (2102.36, 2232.58) | 0.85 (0.84, 0.86) |
| *P*-trend |  | <0.001 | 0.005 | <0.001 |
| **Self-reported Health Status** |  |  |  |  |
| Good or fair | 313 (38.45) | 169.42 (156.75, 182.09) | 2130.20 (2056.88, 2203.52) | 0.85 (0.84, 0.86) |
| Excellent or very good | 501 (61.55) | 142.88 (132.87, 152.90) | 2300.63 (2242.68, 2358.58) | 0.88 (0.87, 0.89) |
| *P*-trend |  | 0.001 | 0.0004 | <0.0001 |
| **Number of multi-morbidities (0-13)\*\*\*** |  |  |  |  |
| None (reference) | 134 (16.67) | 139.70 (120.31, 159.08) | 2264.72 (2152.53, 2376.90) | 0.88 (0.86, 0.89) |
| 1 | 284 (35.32) | 157.74 (144.42, 171.05) | 2271.50 (2194.43, 2348.57) | 0.87 (0.85, 0.88) |
| 2 | 246 (30.60) | 155.04 (140.73, 169.35) | 2217.21 (2134.41, 2300.01) | 0.86 (0.85, 0.87) |
| 3+ | 140 (17.41) | 146.95 (127.97, 165.93) | 2181.69 (2071.84, 2291.54) | 0.87 (0.85, 0.89) |
| *P*-trend |  | 0.76 | 0.18 | 0.47 |
| **Smoking Status** |  |  |  |  |
| Never smoked | 457 (56.07) | 151.87 (141.32, 162.42) | 2241.99 (2180.86, 2303.12) | 0.87 (0.86, 0.88) |
| Current or past smoker | 358 (43.93) | 154.08 (142.16, 166.01) | 2230.16 (2161.07, 2299.24) | 0.86 (0.85, 0.87) |
| *P*-trend |  | 0.79 | 0.80 | 0.60 |

All models are adjusted by clinic site (RAR parameter~clinic site + one descriptive characteristic in separate models)
L5: Avg activity of the 5 consecutive hours with least activity
M10: Avg activity of the 10 consecutive hours with most activity
For predictors with >2 categories, a p-trend was calculated, looking for a linear trend across the categories.
Categories were also compared to the reference category.
The symbols represent the p-value for the comparison of this category to the reference category
Symbols: \*= p-value<0.05; \*\*= p-value<0.01; \*\*\*= p-value<0.001
\*\*\* Mutimorbidity index includes arthritis, cancer (excluding nonmelanoma skin cancer), atrial fibrillation, chronic kidney disease, chronic obstructive pulmonary disease, heart disease, congestive heart failure, dementia, depression, diabetes mellitus, osteoporosis, stroke and aortic stenosis

**Supplemental Table 1B:**
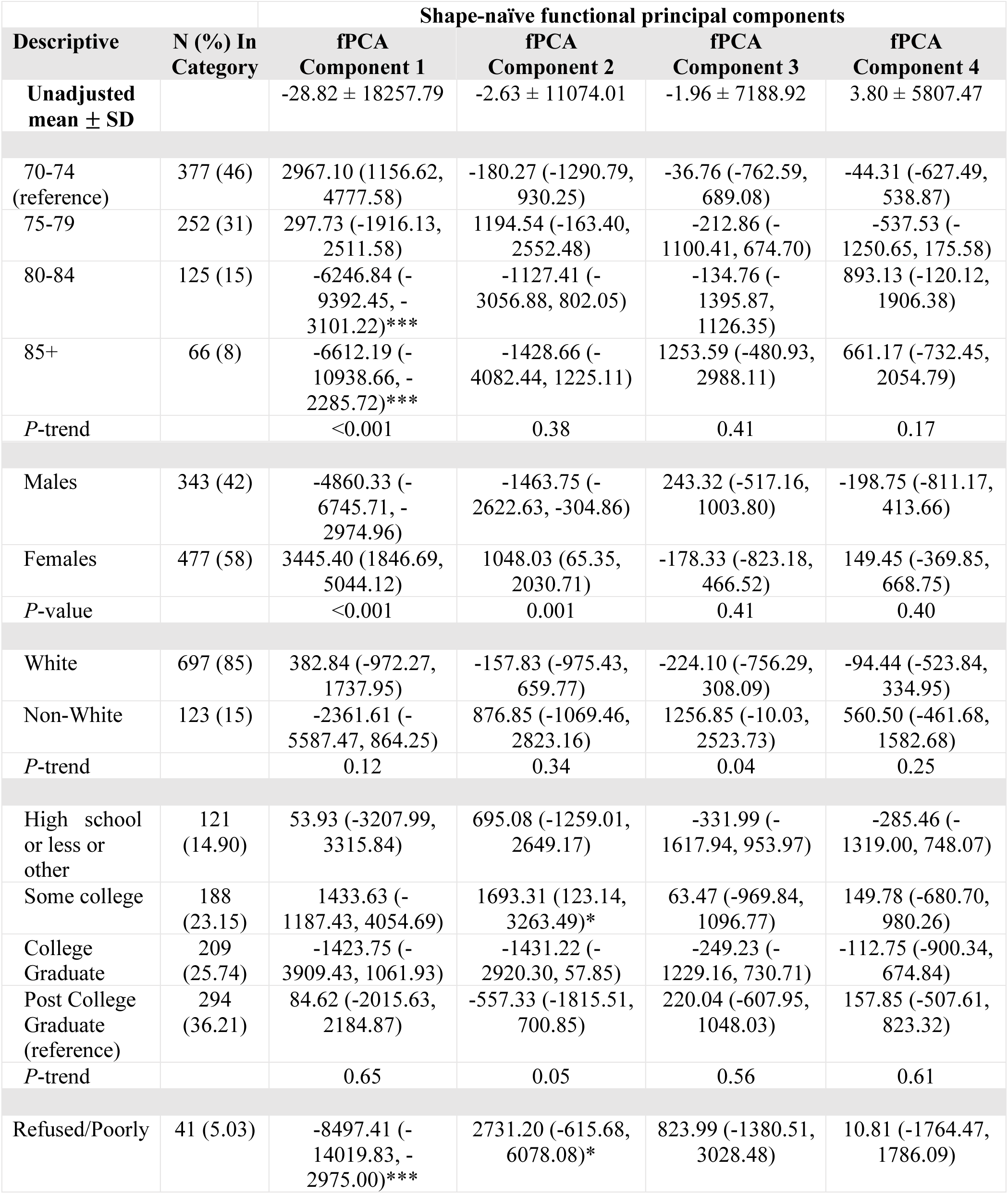
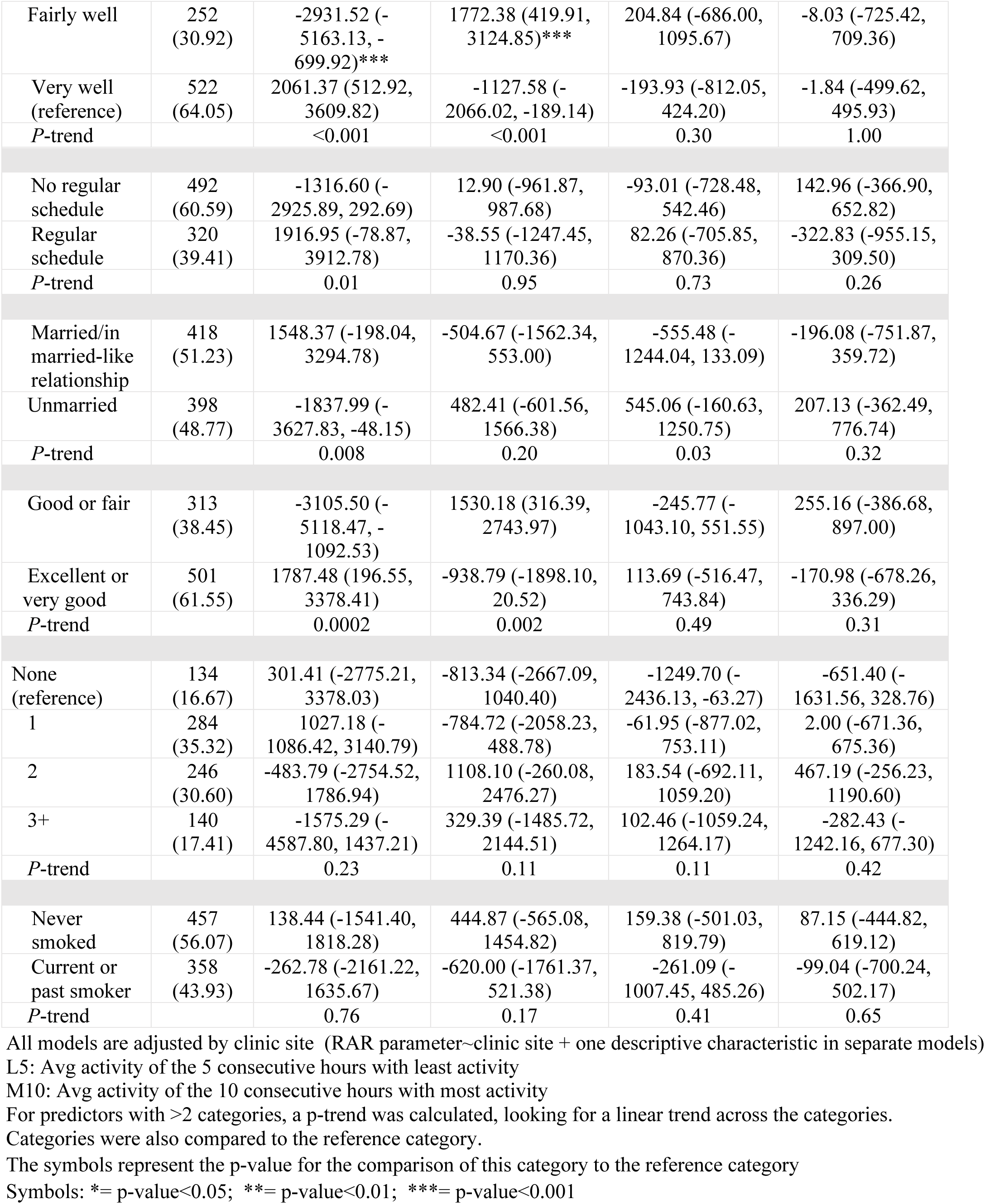
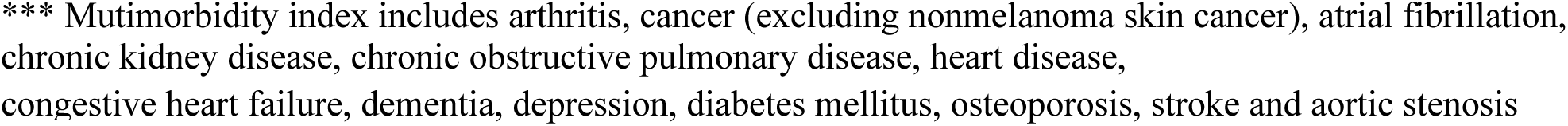
Associations of descriptive variables with rest-activity rhythm parameters. Site adjusted means (95% CI).

**Supplemental Table 2A:** Associations of descriptive variables with parametric rest-activity rhythm parameters. Multivariable adjusted means (95% CI).

|  | Parametric |  |  |  |
| --- | --- | --- | --- | --- |
| Descriptive | Amplitude,<br>counts/min | Mesor,<br>counts/min | Acrophase,<br>portions of hours | Pseudo f-value |
| **Age (yrs)** |  |  |  |  |
| 70-74 (reference) | 2311.28 (2191.99,<br>2430.56) | 1368.73 (1303.89,<br>1433.58) | 14.30 (14.15,<br>14.46) | 664.66 (634.14,<br>695.18) |
| 75-79 | 2183.72 (2040.11,<br>2327.33) | 1294.29 (1216.22,<br>1372.36) | 14.39 (14.21,<br>14.58) | 696.09 (659.35,<br>732.83) |
| 80-84 | 2018.41 (1813.55,<br>2223.27)* | 1228.77 (1117.40,<br>1340.14)* | 14.13 (13.87,<br>14.40) | 625.00 (572.59,<br>677.42) |
| 85+ | 1860.60 (1575.95,<br>2145.26)** | 1185.86 (1031.11,<br>1340.60)* | 14.27 (13.90,<br>14.64) | 689.46 (616.63,<br>762.29) |
| *P*-trend | <0.001 | 0.007 | 0.54 | 0.88 |
| **Sex** |  |  |  |  |
| Males | 2063.09 (1934.14,<br>2192.03) | 1257.77 (1187.67,<br>1327.87) | 14.27 (14.10,<br>14.44) | 580.85 (547.75,<br>613.94) |
| Females | 2282.50 (2174.69,<br>2390.31) | 1346.85 (1288.24,<br>1405.45) | 14.32 (14.18,<br>14.46) | 734.72 (707.04,<br>762.39) |
| *P*-value | 0.01 | 0.07 | 0.66 | <0.001 |
| **Race** |  |  |  |  |
| White | 2215.51 (2129.55,<br>2301.46) | 1314.33 (1267.59,<br>1361.06) | 14.30 (14.18,<br>14.41) | 675.52 (653.46,<br>697.59) |
| Non-White | 2042.52 (1827.28,<br>2257.77) | 1281.26 (1164.25,<br>1398.28) | 14.33 (14.05,<br>14.61) | 639.49 (584.23,<br>694.74) |
| *P*-trend | 0.15 | 0.61 | 0.81 | 0.24 |
| **Education Level** |  |  |  |  |
| High school or<br>less or other | 2319.81 (2108.88,<br>2530.75) | 1397.11 (1282.52,<br>1511.70) | 14.25 (13.97,<br>14.52) | 721.73 (667.56,<br>775.90)** |
| Some college | 2158.88 (1989.53,<br>2328.22) | 1270.33 (1178.33,<br>1362.32) | 14.43 (14.21,<br>14.65) | 705.53 (662.04,<br>749.02)* |
| College Graduate | 2158.26 (2000.35,<br>2316.18) | 1315.31 (1229.53,<br>1401.10) | 14.14 (13.94,<br>14.35) | 653.38 (612.83,<br>693.93) |
| Post College<br>Graduate<br>(reference) | 2181.92 (2047.42,<br>2316.41) | 1294.98 (1221.91,<br>1368.04) | 14.36 (14.18,<br>14.53) | 639.32 (604.78,<br>673.85) |
| *P*-trend | 0.45 | 0.35 | 0.86 | 0.004 |
| **How well money takes care of needs at end of month** |  |  |  |  |
| Refused/Poorly | 2074.24 (1689.09,<br>2459.40) | 1262.09 (1052.73,<br>1471.46) | 14.65 (14.15,<br>15.14) | 574.27 (475.43,<br>673.12)* |
| Fairly well | 2166.52 (2018.09,<br>2314.94) | 1315.85 (1235.17,<br>1396.53) | 14.44 (14.25,<br>14.63) | 619.47 (581.38,<br>657.57)*** |
| Very well<br>(reference) | 2210.23 (2108.98,<br>2311.47) | 1309.87 (1254.83,<br>1364.90) | 14.21 (14.08,<br>14.34) | 701.14 (675.16,<br>727.12) |
| *P*-trend | 0.47 | 0.86 | 0.02 | <0.001 |
| **Work or Volunteer Schedule** |  |  |  |  |
| No regular<br>schedule | 2136.10 (2033.40,<br>2238.80) | 1290.59 (1234.76,<br>1346.42) | 14.30 (14.17,<br>14.44) | 670.87 (644.51,<br>697.23) |
| Regular schedule | 2273.38 (2146.65, 2400.10) | 1338.33 (1269.44, 1407.22) | 14.30 (14.13, 14.46) | 669.59 (637.06, 702.12) |
| *P*-trend | 0.10 | 0.30 | 0.97 | 0.95 |
| **Martial Status** |  |  |  |  |
| Married/in married-like relationship | 2295.88 (2179.50, 2412.27) | 1364.08 (1300.81, 1427.35) | 14.23 (14.08, 14.38) | 713.25 (683.38, 743.12) |
| Unmarried | 2077.55 (1956.32, 2198.79) | 1250.95 (1185.04, 1316.85) | 14.38 (14.22, 14.53) | 624.20 (593.08, 655.32) |
| *P*-trend | 0.02 | 0.02 | 0.23 | <0.001 |
| **Self-reported Health Status** |  |  |  |  |
| Good or fair | 2113.41 (1978.56, 2248.26) | 1264.65 (1191.34, 1337.96) | 14.41 (14.23, 14.58) | 666.58 (631.97, 701.20) |
| Excellent or very good | 2238.17 (2134.47, 2341.87) | 1337.16 (1280.78, 1393.54) | 14.23 (14.10, 14.37) | 672.68 (646.06, 699.30) |
| *P*-trend | 0.17 | 0.14 | 0.13 | 0.79 |
| **Number of multimorbidities (0-13)\*\*\*** |  |  |  |  |
| None (reference) | 2222.83 (2023.03, 2422.62) | 1333.30 (1224.67, 1441.92) | 14.21 (13.95, 14.47) | 716.01 (664.79, 767.23) |
| 1 | 2240.31 (2105.83, 2374.78) | 1331.98 (1258.86, 1405.09) | 14.22 (14.05, 14.40) | 670.58 (636.10, 705.06) |
| 2 | 2160.80 (2016.38, 2305.22) | 1291.40 (1212.88, 1369.92) | 14.44 (14.26, 14.63) | 647.18 (610.16, 684.21)* |
| 3+ | 2111.83 (1915.62, 2308.03) | 1273.48 (1166.80, 1380.16) | 14.30 (14.04, 14.55) | 667.54 (617.24, 717.84) |
| *P*-trend | 0.31 | 0.33 | 0.28 | 0.14 |
| **Smoking Status** |  |  |  |  |
| Never smoked | 2163.29 (2056.01, 2270.57) | 1291.96 (1233.64, 1350.28) | 14.35 (14.22, 14.49) | 659.94 (632.40, 687.47) |
| Current or past smoker | 2225.15 (2104.68, 2345.62) | 1331.71 (1266.22, 1397.20) | 14.24 (14.08, 14.39) | 683.43 (652.51, 714.36) |
| *P*-trend | 0.46 | 0.38 | 0.27 | 0.27 |

One model, adjusted by clinic site and all descriptive visted
For predictors with >2 categories, a p-trend was calculated, looking for a linear trend across the categories.
Categories were also compared to the reference category.
The symbols represent the p-value for the comparison of this category to the reference category.
Symbols: \* = p-value<0.05; \*\* = p-value<0.01; \*\*\* = p-value<0.001
\*\*\* Mutimorbidity index includes arthritis, cancer (excluding nonmelanoma skin cancer), atrial fibrillation, chronic kidney disease, chronic obstructive pulmonary disease, heart congestive heart failure, dementia, depression, diabetes mellitus, osteoporosis, stroke and aortic stenosis

**Supplemental Table 2B:** Associations of descriptive variables with parametric rest-activity rhythm parameters. Multivariable adjusted means (95% CI).

|  | **Nonparametric** |  |  |  |  |
| --- | --- | --- | --- | --- | --- |
| **Descriptive** | **Interdaily stability**<br>**(range 0-1)** | **Intradaily variability**<br>**(range 0-2)** | **L5:**<br>**counts/min** | **M10:**<br>**counts/min** | **Relative amplitude**<br>**((M10-L5)/(M10+L5))** |
| **Age (yrs)** |  |  |  |  |  |
| 70-74<br>(reference) | 0.57 (0.56, 0.58) | 0.88 (0.86, 0.91) | 158.89 (147.23, 170.56) | 2332.11 (2268.07, 2396.15) | 0.87 (0.86, 0.87) |
| 75-79 | 0.59 (0.57, 0.60) | 0.86 (0.84, 0.89) | 146.36 (132.32, 160.40) | 2265.65 (2188.54, 2342.75) | 0.87 (0.86, 0.88) |
| 80-84 | 0.57 (0.55, 0.59) | 0.97 (0.93, 1.01)*** | 136.80 (116.77, 156.84) | 1980.20 (1870.21, 2090.19)*** | 0.86 (0.85, 0.88) |
| 85+ | 0.60 (0.57, 0.63) | 0.94 (0.88, 0.99) | 164.04 (136.21, 191.88) | 2083.66 (1930.83, 2236.50)** | 0.86 (0.84, 0.88) |
| *P*-trend | 0.17 | 0.002 | 0.37 | <0.001 | 0.87 |
| **Sex** |  |  |  |  |  |
| Males | 0.55 (0.54, 0.56) | 0.92 (0.90, 0.95) | 157.48 (144.84, 170.12) | 2003.52 (1933.94, 2073.09) | 0.85 (0.84, 0.86) |
| Females | 0.60 (0.59, 0.61) | 0.87 (0.85, 0.90) | 148.19 (137.62, 158.76) | 2406.38 (2348.21, 2464.55) | 0.88 (0.87, 0.89) |
| *P*-value | <0.001 | 0.005 | 0.29 | <0.001 | <0.001 |
| **Race** |  |  |  |  |  |
| White | 0.58 (0.58, 0.59) | 0.90 (0.88, 0.92) | 145.90 (137.47, 154.32) | 2247.81 (2201.43, 2294.19) | 0.87 (0.87, 0.88) |
| Non-White | 0.55 (0.53, 0.57) | 0.87 (0.82, 0.91) | 189.04 (167.93, 210.14) | 2178.48 (2062.34, 2294.62) | 0.83 (0.82, 0.85) |
| *P*-trend | 0.01 | 0.13 | <0.001 | 0.28 | <0.001 |
| **Education Level** |  |  |  |  |  |
| High school or less or other | 0.58 (0.56, 0.61) | 0.86 (0.82, 0.90)** | 169.24 (148.56, 189.92) | 2336.96 (2223.14, 2450.78)* | 0.86 (0.84, 0.87) |
| Some college | 0.59 (0.57, 0.61) | 0.85 (0.82, 0.89)*** | 151.25 (134.64, 167.85) | 2285.17 (2193.80, 2376.55) | 0.87 (0.86, 0.89) |
| College Graduate | 0.57 (0.56, 0.59) | 0.92 (0.89, 0.95) | 151.54 (136.06, 167.03) | 2190.20 (2104.99, 2275.40) | 0.86 (0.85, 0.87) |
| Post College Graduate (reference) | 0.58 (0.56, 0.59) | 0.92 (0.89, 0.95) | 146.02 (132.83, 159.21) | 2201.78 (2129.21, 2274.35) | 0.87 (0.86, 0.88) |
| *P*-trend | 0.41 | 0.001 | 0.11 | 0.03 | 0.44 |
| **How well money takes care of needs at end of month** |  |  |  |  |  |
| Refused/Poorly | 0.54 (0.50, 0.58)** | 0.94 (0.87, 1.02) | 172.08 (134.35, 209.81) | 2009.18 (1801.37, 2217.00)** | 0.84 (0.81, 0.87)* |
| Fairly well | 0.56 (0.54, 0.57)*** | 0.90 (0.87, 0.93) | 170.06 (155.52, 184.61)** | 2140.39 (2060.31, 2220.48)*** | 0.85 (0.84, 0.86)*** |
| Very well (reference) | 0.59 (0.58, 0.60) | 0.89 (0.87, 0.91) | 142.15 (132.23, 152.07) | 2299.90 (2245.27, 2354.53) | 0.88 (0.87, 0.89) |
| *P*-trend | <0.001 | 0.23 | 0.003 | <0.001 | <0.001 |
| **Work or Volunteer Schedule** |  |  |  |  |  |
| No regular schedule | 0.59 (0.57, 0.60) | 0.90 (0.88, 0.92) | 147.54 (137.48, 157.61) | 2218.00 (2162.58, 2273.41) | 0.87 (0.86, 0.88) |
| Regular schedule | 0.57 (0.56, 0.59) | 0.88 (0.86, 0.91) | 158.93 (146.51, 171.35) | 2267.96 (2199.58, 2336.33) | 0.86 (0.86, 0.87) |
| *P*-trend | 0.13 | 0.20 | 0.17 | 0.27 | 0.49 |
| **Martial Status** |  |  |  |  |  |
| Married/in married-like relationship | 0.60 (0.59, 0.61) | 0.90 (0.88, 0.92) | 138.17 (126.76, 149.58) | 2322.14 (2259.35, 2384.94) | 0.88 (0.88, 0.89) |
| Unmarried | 0.56 (0.54, 0.57) | 0.89 (0.87, 0.91) | 167.04 (155.16, 178.93) | 2147.19 (2081.77, 2212.60) | 0.85 (0.84, 0.86) |
| *P*-trend | <0.001 | 0.60 | 0.001 | <0.001 | <0.001 |
| **Self-reported Health Status** |  |  |  |  |  |
| Good or fair | 0.58 (0.56, 0.59) | 0.90 (0.87, 0.92) | 158.39 (145.17, 171.61) | 2172.92 (2100.16, 2245.68) | 0.86 (0.85, 0.87) |
| Excellent or very good | 0.58 (0.57, 0.59) | 0.89 (0.87, 0.91) | 148.20 (138.04, 158.37) | 2277.73 (2221.78, 2333.69) | 0.87 (0.87, 0.88) |
| *P*-trend | 0.82 | 0.72 | 0.25 | 0.03 | 0.02 |
| **Number of multimorbidities (0-13)\*\*\*** |  |  |  |  |  |
| None (reference) | 0.60 (0.57, 0.62) | 0.87 (0.83, 0.91) | 147.75 (128.19, 167.32) | 2277.13 (2169.30, 2384.95) | 0.87 (0.86, 0.89) |
| 1 | 0.57 (0.56, 0.59) | 0.91 (0.89, 0.94) | 160.03 (146.86, 173.19) | 2253.07 (2180.50, 2325.65) | 0.86 (0.85, 0.87) |
| 2 | 0.58 (0.56, 0.59) | 0.89 (0.86, 0.92) | 150.39 (136.24, 164.53) | 2219.91 (2141.97, 2297.85) | 0.87 (0.86, 0.88) |
| 3+ | 0.58 (0.56, 0.60) | 0.90 (0.86, 0.93) | 142.91 (123.70, 162.13) | 2201.33 (2095.44, 2307.21) | 0.87 (0.86, 0.89) |
| *P*-trend | 0.69 | 0.71 | 0.47 | 0.27 | 0.69 |
| **Smoking Status** |  |  |  |  |  |
| Never smoked | 0.58 (0.56, 0.59) | 0.90 (0.88, 0.92) | 150.18 (139.66, 160.69) | 2224.43 (2166.55, 2282.32) | 0.87 (0.86, 0.88) |
| Current or past smoker | 0.59 (0.57, 0.60) | 0.89 (0.86, 0.91) | 154.46 (142.65, 166.27) | 2254.75 (2189.75, 2319.75) | 0.87 (0.86, 0.88) |
| *P*-trend | 0.19 | 0.39 | 0.60 | 0.50 | 0.71 |

One model, adjusted by clinic site and all descriptive visted.
L5: Avg activity of the 5 consecutive hours with least activity
M10: Avg activity of the 10 consecutive hours with most activity
For predictors with >2 categories, a p-trend was calculated, looking for a linear trend across the categories.
Categories were also compared to the reference category.
The symbols represent the p-value for the comparison of this category to the reference category.
Symbols: \* = p-value<0.05; \*\* = p-value<0.01; \*\*\* = p-value<0.001
\*\*\* Mutimorbidity index includes arthritis, cancer (excluding nonmelanoma skin cancer), atrial fibrillation, chronic kidney disease, chronic obstructive pulmonary disease, heart congestive heart failure, dementia, depression, diabetes mellitus, osteoporosis, stroke and aortic stenosis

**Supplemental Table 2C:** Associations of descriptive variables with fPCA parameters. Multivariable adjusted means (95% CI).

|  | Shape-naïve functional principal components |  |  |  |
| --- | --- | --- | --- | --- |
| Descriptive | fPCA Component 1 | fPCA Component 2 | fPCA Component 3 | fPCA Component 4 |
| **Age (yrs)** |  |  |  |  |
| 70-74<br>(reference) | 2256.35 (497.16,<br>4015.54) | -166.11 (-1301.35,<br>969.13) | -305.62 (-1042.40,<br>431.17) | -121.13 (-728.43,<br>486.17) |
| 75-79 | 621.80 (-1496.14,<br>2739.74) | 1348.26 (-18.48,<br>2715.01) | -219.81 (-1106.85,<br>667.23) | -566.13 (-1297.28,<br>165.02) |
| 80-84 | -6268.84 (-9290.14, -<br>3247.53)*** | -1455.78 (-3405.49,<br>493.93) | -335.64 (-1601.03,<br>929.75) | 836.49 (-206.52,<br>1879.49) |
| 85+ | -3875.73 (-8073.82,<br>322.36)** | -1462.68 (-4171.79,<br>1246.43) | 880.15 (-878.11,<br>2638.40) | 662.27 (-786.97,<br>2111.52) |
| P-trend | <0.001 | 0.31 | 0.41 | 0.16 |
| **Sex** |  |  |  |  |
| Males | -6568.04 (-8476.85, -<br>4659.23) | -1167.27 (-2399.17,<br>64.63) | 172.23 (-624.66,<br>969.12) | -248.97 (-906.98,<br>409.03) |
| Females | 4647.95 (3051.99,<br>6243.90) | 840.75 (-189.25,<br>1870.74) | -446.06 (-1112.34,<br>220.22) | 93.82 (-456.34,<br>643.98) |
| P-value | <0.001 | 0.02 | 0.26 | 0.45 |
| **Race** |  |  |  |  |
| White | 187.44 (-1085.04,<br>1459.92) | 41.65 (-779.58,<br>862.88) | -356.90 (-888.14,<br>174.33) | -155.79 (-594.44,<br>282.86) |
| Non-White | -1422.58 (-4608.94,<br>1763.78) | -242.93 (-2299.33,<br>1813.48) | 826.17 (-504.07,<br>2156.42) | 585.95 (-512.45,<br>1684.36) |
| P-trend | 0.36 | 0.80 | 0.11 | 0.22 |
| **Education Level** |  |  |  |  |
| High school or<br>less or other | 2251.14 (-871.33,<br>5373.61) | 622.63 (-1391.14,<br>2636.40) | -737.37 (-2041.76,<br>567.03) | -464.99 (-1541.98,<br>612.00) |
| Some college | 1183.77 (-1323.04,<br>3690.58) | 855.06 (-761.65,<br>2471.78) | -200.60 (-1247.80,<br>846.61) | -18.99 (-883.63,<br>845.65) |
| College<br>Graduate | -1364.14 (-3701.68,<br>973.39) | -1028.23 (-2535.77,<br>479.31) | -317.21 (-1293.70,<br>659.28) | -191.11 (-997.36,<br>615.14) |
| Post College<br>Graduate<br>(reference) | -807.60 (-2798.51,<br>1183.31) | -57.19 (-1341.18,<br>1226.81) | 136.93 (-694.76,<br>968.63) | 200.96 (-485.74,<br>887.65) |
| P-trend | 0.07 | 0.39 | 0.31 | 0.37 |
| **How well money takes care of needs at end of month** |  |  |  |  |
| Refused/Poorly | -6415.87 (-12116.41, -<br>715.33)** | 1262.82 (-2414.25,<br>4939.88) | 576.48 (-1803.46,<br>2956.42) | -243.63 (-2209.11,<br>1721.85) |
| Fairly well | -3273.47 (-5470.32, -<br>1076.63)*** | 1497.26 (80.21,<br>2914.30)* | -219.65 (-1136.82,<br>697.52) | -178.51 (-935.95,<br>578.94) |
| Very well<br>(reference) | 1929.48 (430.98,<br>3427.98) | -797.12 (-1763.71,<br>169.47) | -224.48 (-850.10,<br>401.14) | 25.06 (-491.61,<br>541.73) |
| P-trend | <0.001 | 0.02 | 0.70 | 0.66 |
| **Work or Volunteer Schedule** |  |  |  |  |
| No regular<br>schedule | -670.62 (-2190.94,<br>849.70) | -1.01 (-982.19,<br>980.17) | -340.09 (-974.79,<br>294.62) | 136.29 (-387.80,<br>660.37) |
| Regular schedule | 906.05 (-969.90, 2781.99) | 3.77 (-1206.92, 1214.47) | 43.41 (-739.76, 826.59) | -330.68 (-977.36, 315.99) |
| *P*-trend | 0.20 | 1.00 | 0.46 | 0.28 |
| **Martial Status** |  |  |  |  |
| Married/in married-like relationship | 2237.75 (514.89, 3960.60) | 179.32 (-932.57, 1291.21) | -686.66 (-1405.92, 32.60) | -79.24 (-673.14, 514.67) |
| Unmarried | -2498.11 (-4292.79, -703.44) | -191.15 (-1349.40, 967.09) | 349.83 (-399.41, 1099.07) | -17.61 (-636.27, 601.05) |
| *P*-trend | <0.001 | 0.67 | 0.06 | 0.89 |
| **Self-reported Health Status** |  |  |  |  |
| Good or fair | -1915.80 (-3912.03, 80.44) | 1063.68 (-224.65, 2352.00) | -660.93 (-1494.31, 172.46) | 152.29 (-535.85, 840.43) |
| Excellent or very good | 1105.71 (-429.42, 2640.85) | -651.12 (-1641.86, 339.62) | 103.00 (-537.88, 743.89) | -173.39 (-702.58, 355.81) |
| *P*-trend | 0.02 | 0.05 | 0.17 | 0.48 |
| **Number of multimorbidities (0-13)\*\*\*** |  |  |  |  |
| None (reference) | 586.97 (-2371.12, 3545.06) | -159.37 (-2065.94, 1747.21) | -1328.10 (-2562.26, -93.95) | -654.70 (-1672.16, 362.76) |
| 1 | 444.25 (-1546.75, 2435.24) | -640.63 (-1923.88, 642.62) | -149.79 (-980.46, 680.88) | 6.71 (-678.11, 691.52) |
| 2 | -364.98 (-2503.20, 1773.23) | 888.70 (-489.44, 2266.84) | 113.62 (-778.47, 1005.72) | 443.19 (-292.27, 1178.64) |
| 3+ | -1068.71 (-3973.70, 1836.27) | -104.13 (-1976.48, 1768.21) | 286.05 (-925.95, 1498.04) | -460.71 (-1459.90, 538.48) |
| *P*-trend | 0.36 | 0.50 | 0.08 | 0.60 |
| **Smoking Status** |  |  |  |  |
| Never smoked | -366.11 (-1954.20, 1221.98) | 544.94 (-479.98, 1569.86) | -77.97 (-740.97, 585.02) | 125.81 (-421.64, 673.25) |
| Current or past smoker | 361.94 (-1421.37, 2145.24) | -681.50 (-1832.41, 469.41) | -324.79 (-1069.29, 419.70) | -269.51 (-884.25, 345.24) |
| *P*-trend | 0.55 | 0.12 | 0.63 | 0.35 |

One model, adjusted by clinic site and all descriptive variables listed.
For predictors with >2 categories, a *p*-trend was calculated, looking for a linear trend across the categories. Categories were also compared to the reference category.
The symbols represent the *p*-value for the comparison of this category to the reference category
Symbols: \*= *p*-value<0.05; \*\*= *p*-value<0.01; \*\*\*= *p*-value<0.001
\*\*\* Mutimorbidity index includes arthritis, cancer (excluding nonmelanoma skin cancer), atrial fibrillation, chronic kidney disease, chronic obstructive pulmonary disease, heart disease, congestive heart failure, dementia, depression, diabetes mellitus, osteoporosis, stroke and aortic stenosis.

**Supplemental Figure 1.**
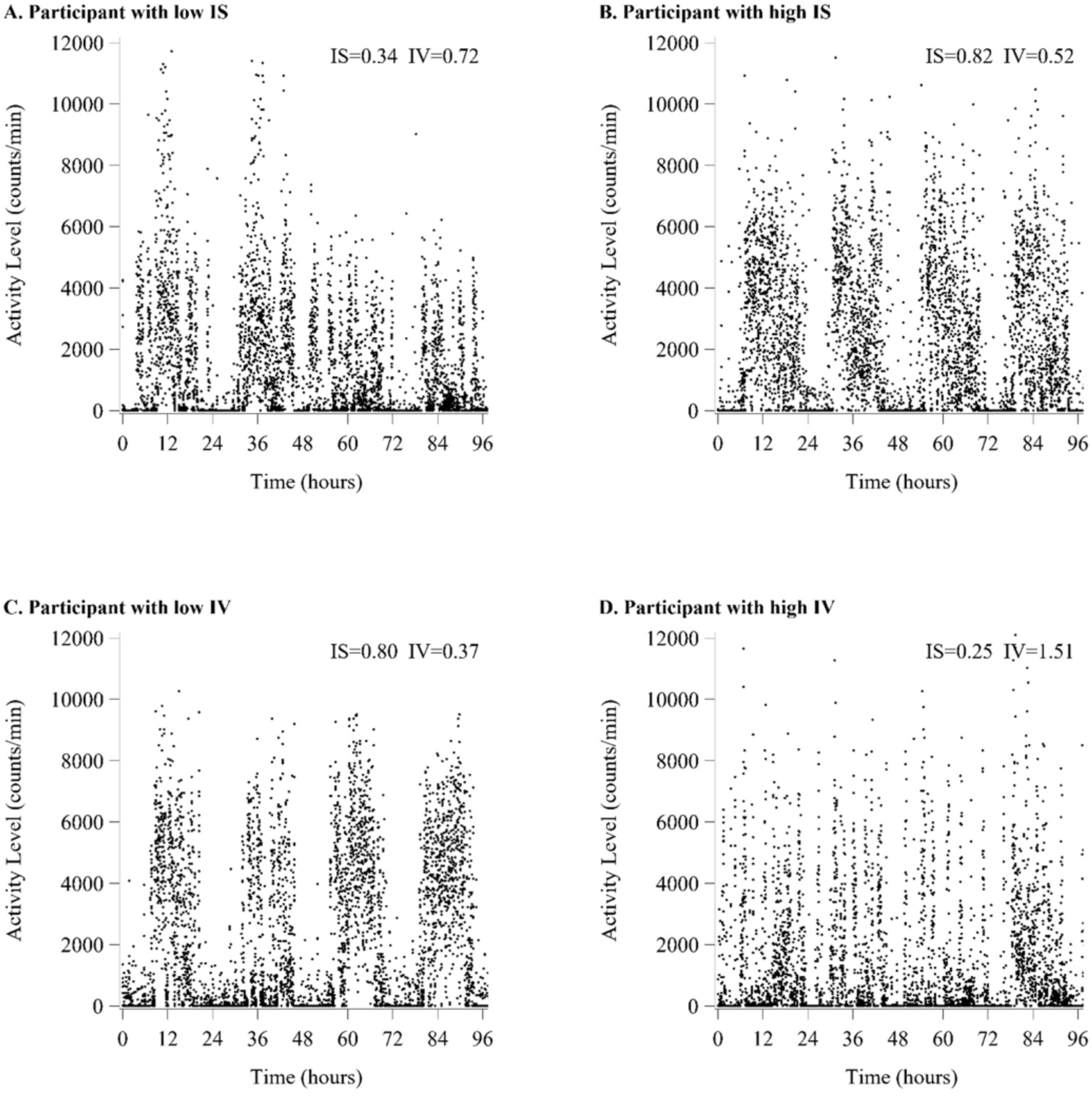
Graphical representations of low IS vs high IS, as well as low IV vs. high IV, from rest-activity rhythm profiles. Comparison of representative rest-activity rhythm plots of individual participants from the lowest 10^th^ percentile IS (Panel A) versus highest 10^th^ percentile IS (Panel B). Comparison of representative rest-activity rhythms of individual participants from the lowest decile IV (Panel C) versus the highest decile values for IV (Panel D) to graphically show variance in rhythmic stability across the 8-day period. Each plot point represents an activity count.

**Supplemental Figure 2.**
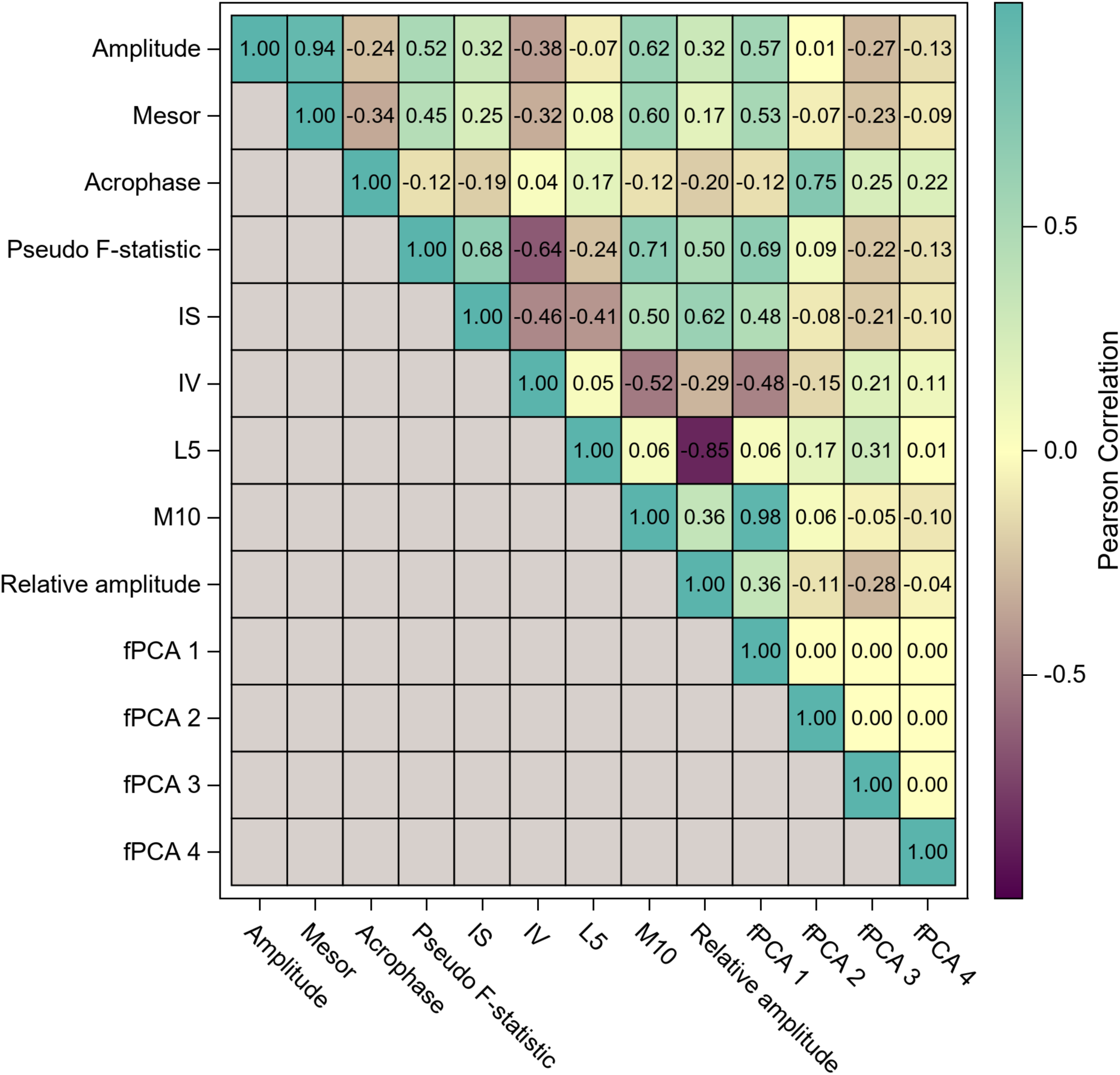
Unadjusted correlations of rest-activity rhythm parameters. Correlations matrix between RAR rhythms, revealing generally good agreement between parametric vs. non-parametric variables.

